# The patient experience of COVID-19: A qualitative investigation with symptomatic outpatients

**DOI:** 10.1101/2021.06.02.21257739

**Authors:** Diana Rofail, Nadine McGale, Anna J. Podolanczuk, Alissa Rams, Krystian Przydzial, Sumathi Sivapalasingam, Vera Mastey, Patrick Marquis

## Abstract

**Background:** Coronavirus disease 2019 (COVID-19) is an acute respiratory illness characterised by a range of symptoms. Severe cases of COVID-19 could lead to hospitalisation and intensive care unit admission. However, little is known about the symptomatic experience of outpatients with COVID-19, or its daily life impact; the objective of this qualitative research was to document this experience.

**Methods:** Thirty US adult patients with COVID-19 were interviewed within 21 days of diagnosis. To be included, patients had to self-report one of the following: fever, cough, shortness of breath/difficulty breathing, change/loss of taste/smell, vomiting/diarrhoea or body/muscle aches. Patients were asked open-ended questions to elicit their symptoms and the impacts COVID-19 had on their daily lives. Interviews were transcribed and inductively coded to perform thematic analysis and propose a conceptual model. The adequacy of the sample size was assessed by conceptual saturation analysis. Five independent clinicians were also interviewed about their experience treating outpatients with COVID-19.

**Findings:** Patient-reported concepts were organised into six symptom themes (upper respiratory, lower respiratory, systemic, gastrointestinal, smell and taste, and other) and seven impact themes (activities of daily living, broad daily activities, leisure/social activities, physical impacts, emotional impacts, professional impacts and quarantine-specific impacts). Symptom type, severity, duration and time of onset varied greatly by patient. Clinicians endorsed all symptoms reported by patients.

**Interpretation:** The manifestation of symptoms in outpatient COVID-19 is heterogeneous and affects all aspects of daily life. While reported symptoms were in line with previously published reports, patients offered new detailed insights into their symptomatic experiences and the impacts that symptoms had on their daily lives. Future studies should explore the symptoms and impacts of COVID-19 longitudinally, to better understand their early onset, their progression/resolution and the possible long-term implications of COVID-19.

**Funding:** This research was sponsored and paid for by Regeneron Pharmaceuticals, Inc.

**Research in context:** *Evidence before this study:* Databases used: PubMed. Criteria for inclusion: qualitative study of the outpatient experience of COVID-19 (December 2019 – March 2021). Search terms included “qualitative” OR “interviews” AND “COVID-19” or “coronavirus”. The quality of the evidence was deemed not relevant to the current study, as the work was related to other contexts (eg, hospitalized patients or treatment of COVID-19), or data was collected quantitively (ie, questionnaire/survey) and did not include an in-depth perspective of the outpatient experience.

*Added value of this study:* This research fills important gaps in the literature, as the outpatient experience (ie, symptoms and daily life impacts) of COVID-19 has not been documented qualitatively.

*Implications of all the available evidence:* Findings of this research may be used to supplement existing knowledge of the outpatient experience of mild-to-moderate COVID-19, to further inform treatment guidelines. This work also provides an evidence base for identifying the symptoms and impacts important for evaluating potential treatment benefit in outpatients.

## Introduction

Coronavirus disease 2019 (COVID-19) is a disease caused by severe acute respiratory syndrome coronavirus 2 (SARS-CoV-2), a novel coronavirus, that the World Health Organization declared a pandemic in March 2020.^1^ Since then there have been over 163 million documented cases and over 3 million confirmed deaths in 192 countries, as of May 2021.^2^ Initial reports of common COVID-19 symptoms included fever, cough and shortness of breath.^3^ By May 2020, the Centers for Disease Control (CDC) had updated its guidelines with additional symptoms: loss of smell/taste, chills, fatigue, sore throat, nasal congestion and gastrointestinal symptoms like vomiting and diarrhoea. Since then, our understanding has evolved regarding longevity of symptoms, incubation period, those at risk of severe illness, and reinfection.^4^ Most of the data used to inform COVID-19 presentation is based on hospitalised patients, and the focus remains on cough, fever and loss of smell/taste as key symptoms. However, recent data from broader samples identified via community testing also confirm fatigue, loss of appetite and chills as predictive of a positive test for SARS-CoV-2.^5^

Little is known about the experience of outpatients with COVID-19, especially how symptoms impact patients’ daily lives (ie, the burden of living with/managing COVID-19, COVID-19’s impact on patients’ activities of daily living, physical functioning and emotional wellbeing). The patients’ voice at the forefront of this research will help us to better understand and adequately address their needs, facilitate dialogue with their healthcare professionals, and support the development of patient-reported outcome instruments for clinical trials^6^ and real-world observational studies. The aim of this qualitative research was to explore the experience of outpatients with COVID-19 based in the US, with regard to symptoms and the impact on daily life.

## Methods

This was a cross-sectional qualitative research study comprising in-depth, open-ended interviews in patients with confirmed diagnosis of COVID-19, complemented by five clinician interviews to integrate a clinical perspective. Interviews were conducted in September and October 2020.

### Participants

Our patient sample was recruited through a healthcare research firm, via healthcare provider networks, COVID-19 support groups, testing centres and social media. The sample comprised thirty adults (aged 18+ years) in the US with a diagnosis of COVID-19, confirmed by positive polymerase chain reaction test within 21 days of interview, who self-reported one of the following: fever (≥100° F, or feeling feverish or chills), cough, shortness of breath or difficulty breathing, change in or loss of taste or smell, vomiting or diarrhoea, or body or muscle aches. Patients were excluded if they were hospitalised in the past 30 days, could not speak English fluently, or currently resided in a nursing home. The target number was 30 patients with the aim of achieving conceptual saturation, ie, the point at which no new information emerges from additional interviews.^7,8^ Five clinicians who regularly treated patients with COVID-19 (≥5 a week) were also interviewed. Recruitment of clinicians was through the same healthcare research firm, with additional support from Regeneron Pharmaceuticals Inc.

### Interview conduct

Patient interviews (∼60 minutes) were conducted virtually by experienced qualitative researchers who received specific training for the current project. A semi-structured interview guide was used to explore the experience of COVID-19, including symptoms and impacts on daily life. First, the interviewer asked open-ended questions; targeted probes were used when patients did not spontaneously detail how the symptom changed from day to day (ie, improved/worsened) or its duration (within-day and across day), severity and/or resolution. Interviewers also used structured prompts for all common symptoms of COVID-19^9^ not spontaneously mentioned. The impacts of symptoms and diagnosis were also explored, including how patients were impacted emotionally and socially (ie, family/friends), as well as any disruption to daily activities (ie, work, hobbies, exercise and chores) due to COVID-19 symptoms.

Clinicians were asked about symptoms observed in outpatients, the clinical presentations and symptomatic profiles of COVID-19, improvement/worsening in symptoms over time, onset/resolution of symptoms, and any clinical hierarchy of symptoms.

### Ethics

Study documents, including the protocol, demographic and health information form, interview guide, screener and informed consent form, received ethical approval from the New England Independent Review Board (study number: 1291666) prior to any contact with patients.

### Data analysis

Interviews were audio-recorded, transcribed and anonymised. A data-driven approach was adopted whereby transcripts were coded in ATLAS.ti software^10^ using an open, inductive, thematic coding approach.^10-13^ The first transcript was independently coded by three researchers; any inconsistencies in codes were discussed and reconciled. Researchers met regularly to discuss and adjust coding guidelines when necessary. Following coding, a conceptual model was developed; this was a visual representation of patient-reported concepts (ie, aspects of their COVID-19 experience) using standard analytical techniques.^10,12,14^ Codes and quotations were compared with the rest of the data (ie, other codes and quotations) and inductively categorised into higher-order overarching categories including themes and sub-themes, reflecting their conceptual underpinning. This involved an iterative process of cross-referencing and comparison between the different analytical categories, which was reviewed and fine-tuned by the research team. The adequacy of the sample size was assessed by saturation analysis^7,8^ performed on quintiles of subsequent transcripts.

## Results

### Patient interviews

All recruited outpatients were interviewed and included in analysis (n=30). Patients were 45.0±19.4 years old (range 18-76), 60% female, and 87% white. The interview was conducted an average of 19 days after symptoms began, and an average of 12 days (median 9.5, range 6–21) after COVID-19 diagnosis. Most patients (83%) were still experiencing at least one symptom during the time of the interview; of those, 68% described the overall severity of their symptoms in the past 24 hours as ‘mild’ and 32% as ‘moderate’. See Table 1 for full patient demographic and health information.

**Table 1:** Overview of patient sample characteristics.

|  | **n=30** |
| --- | --- |
| Age, mean (SD) | 45.03 (19.39) |
| Gender, n (%) |  |
| Male | 12 (40) |
| Female | 18 (60) |
| Race, n (%) |  |
| White | 26 (86.7) |
| Other | 4 (13.3) |
| Number of days between diagnosis and interview, mean (SD) | 12.40 (5.29) |
| Number of days since symptoms began, mean (SD) | 19.17 (7.96) |
| Number of days between symptoms began and diagnosis, mean (SD) | 6.77 (7.96) |
| General health ratings, n (%) |  |
| Excellent | 6 (20) |
| Very good | 8 (26.7) |
| Good | 4 (13.3) |
| Fair | 9 (30) |
| Poor | 3 (10) |
| Co-morbidities, n (%) | 17 (57) |
| SD=standard deviation. |  |

The interviews yielded two overarching domains: symptoms and impacts on daily life. All symptoms reported by patients were identified in the first 25 interviews; the final five interviews yielded three additional impacts on daily life. Thus, conceptual saturation was achieved for reported symptoms.

### Symptoms

Symptoms were categorised into six major themes: upper respiratory tract, lower respiratory tract, systemic, gastrointestinal, smell and taste, and other. Each theme was further broken down into sub-themes, as outlined in Figure 1. Categorisation was created for the purpose of summarising the data, but was not indicative of an underlying common theoretical underpinning.

**Figure 1:**
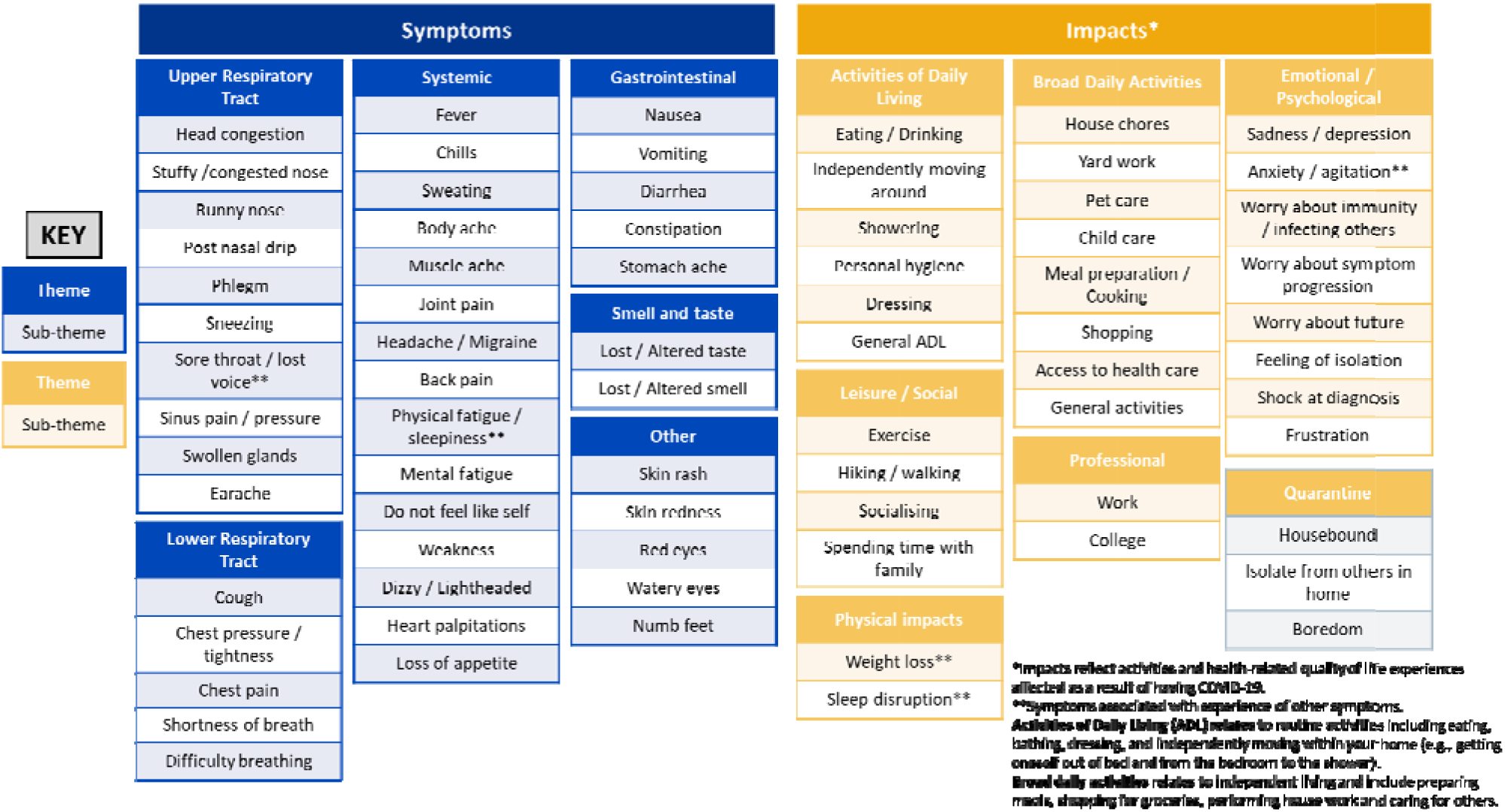
Patient experience of COVID-19.

Upper respiratory tract symptoms included stuffy/congested nose, runny nose, post-nasal drip, phlegm, sneezing, sore throat (accompanied by lost voice), sinus pressure/pain, head congestion, swollen glands and earache. Symptoms were grouped into this theme if they referred to experiences affecting the head, nose or throat. Patients often reported experiencing multiple upper respiratory tract symptoms simultaneously, as one patient stated:

> *“My sinus congestion. I would sneeze a lot, and I was just very stuffy to where my voice didn’t sound normal. It felt like a sinus infection. Most days I had to have a humidifier to try to get it to go away. I still have some stuffiness and runny nose, but it’s a lot better than it was at first.” (Female, 18-24year age range)*

Themes were created based on the way that patients described their symptom experience (eg, patients described stuffy/congested nose differently than head congestion):

> *“It was a little bit of a nasal drip and I was sniffling, and then it developed into what felt like, just complete sinus congestion. No matter what I did, I couldn’t seem to get it cleared out of my system.” (Male, 60-69)*
>
> *“I had head congestion from the very beginning. That was one of the things. When I said I thought I had allergies, it was head congestion. That lasted the entire time, but I felt like I didn’t have mucus in my head that was making me congested. It felt like it was swollen tissues.” (Female, 40-49)*

Lower respiratory tract symptoms were cough, chest pressure/tightness, chest pain, shortness of breath and difficulty breathing (ie, symptoms related to lung functioning). Cough was commonly described as dry and non-productive.

> *“When I cough, it’s coming from my chest really deep. But I haven’t coughed up anything.” (Female, 25-30)*

Similar to upper respiratory symptoms, patients reported multiple lower respiratory symptoms at once (eg, cough and chest pain or chest pressure and difficulty breathing).

> *“I did have that pain because I was coughing a lot. I was trying to take the stuff out of my body, and it kind of hurt a little bit.” (Male, 58)*
>
> *“It just feels like a pressure on my chest where I can’t breathe very well.” (Female, 60-69)*

Gastrointestinal symptoms were nausea, vomiting, diarrhoea, constipation and stomach-ache. These symptoms were not reported as often as others but had a significant impact on the patients’ daily lives.

> *“I actually had started vomiting. It was waking me up at three or four o’clock in the morning.” (Male, 31-39)*
>
> *“I did wake up that morning and I did have a pretty good deal of diarrhoea.” (Male, 70-80)*

The smell and taste theme comprised symptoms related to lost or altered sense of smell or taste. This theme was distinct from upper respiratory tract symptoms because of the pathophysiological mechanisms underlying these symptoms, which are different to that of other common upper-respiratory illnesses where senses are more directly related to other respiratory symptoms (eg, stuffy nose impairing smell).^15^ The experience of changes in smell/taste also differed from other nasal symptoms (ie, stuffed or runny nose). Patients described their sense of taste/smell changing, being diminished or lost altogether. Changes in these senses often occurred together but could occur separately. Lost or altered taste or smell was the only common symptom reported as unchanging while present.

> *“I never completely lost my taste, it remained altered until I began to be able to smell again.” (Female, 18-24)*
>
> *“It’s pretty much stayed the same because I haven’t noticed any highs and lows with it at all, [my smell and taste] just hasn’t been there.” (Male, 31-39)*

The systemic theme reflected symptoms that affect the entire body, rather than a single organ or body part. This theme comprised fever, chills, sweating, body aches, muscle aches, joint pain, headache/migraine, back pain, physical fatigue/sleepiness, mental fatigue, weakness, dizziness/lightheadedness, heart palpitations, loss of appetite and not feeling like yourself. The duration of systemic symptoms was variable; fever, body aches and headache persisted from 1 day to several weeks, which was similarly described in other themes. Fatigue also varied, but typically lasted for at least 1 week (up to 3 weeks for one patient). Varying experiences were also reported in terms of fluctuation of symptoms across all themes but particularly within systemic symptoms, with patients reporting consistency across days, variation within a day (ie, worse in the morning), and variation across days (ie, absent one day but present the next day).

> *“It would change throughout the day. It always seemed that the fevers were worse at night.” (Male, 18-24)*
>
> *“I’m having a good day like, I’m not aching as much, and then, the next day, I would be very sore and achy.” (Female, 60-69)*
>
> *“Some days I don’t have [headache], and other days I do. It’s not very consistent.” (Female, 25-30)*

Other, less common, symptoms reported were skin rash, skin redness, red eyes, watery eyes and numb feet.

### Impacts on daily life

Impacts on daily life referred to any COVID-19 related effects on activities of daily living (ADLs), broad daily activities (eg, day-to-day activities outside of self-care and hygiene), leisure and social life, professional impacts, psychological/emotional impacts, physical impacts and impacts specifically related to quarantine (Figure 1). Most impacts on daily life were not attributed to any singular symptom, but rather to the experience of COVID-19 infection as a whole; though the duration of symptoms was relatively short, patients reported a significant daily life impact.

The ADL theme referred to changes in patients’ performance of self-care tasks (ie, hygiene and dressing), eating habits, mobility or daily routine. Broad daily activities encompassed changes in activities not necessary for fundamental functioning, but which allow patients to live independently, including shopping, meal preparation, home maintenance, yard work and care of children/pets. Each of these activities was reported as central to patients’ lives pre-COVID-19, and patients emphasised the disruption caused by their illness.

> *“I couldn’t get anything done that I normally get [done]. Doing any kind of housework, laundry – I couldn’t do anything.” (Female, 40-49)*
>
> *“[COVID-19] completely interferes with any of your planned activities that would be normal when you’re not sick.” (Male, 60-69)*

Activities related to exercise, hobbies/leisure activities, and socialising with friends or family were categorised under leisure and social life. This theme referred specifically to social/leisure impacts caused by COVID-19 symptoms, not by mandated quarantine due to infection. This theme also included impacts on relationships with those within the patients’ households.

> *“It’s still really, really difficult not being able to hug my kids and my wife and having to stay away from them.” (Male, 31-39)*
>
> *“I’d just say exercise is the main, it’s the worst, like the hardest thing for me to get done.” (Male, 18-24)*

Professional impacts referred to any effects on the patient’s work or schooling. Similarly, this theme comprised only changes due to COVID-19 and its symptoms, not due to quarantine (eg, transition to working from home or virtual class).

> *“I still have school to do all of my online courses and so I would do those for about 3 hours each day and then after that I felt like I just had to be asleep and I just didn’t really have any motivation to do anything else other than watch movies and lay in bed.” (Male, 18-24)*

Physical impacts comprised physical changes related to COVID-19, specifically weight loss and sleep disruption. Though these may be considered symptoms themselves, patients described them as secondary to other symptoms.

> *“I actually lost like 8 pounds during my 2 weeks of quarantine [due to infection].” (Female, 18-24)*
>
> *“I couldn’t get comfortable, and I think that the sleep was disrupted.” (Female, 25-30)*

The emotional/psychological theme encompassed changes in the patients’ psyche or mood, including depression, anxiety, irritability, frustration, feelings of isolation and worries about the symptoms and meaning of a COVID-19 diagnosis. Patients used a breadth of language to describe their emotional response to COVID-19 and its symptoms; sub-themes reflect patients’ words with as much granularity as possible without over-emphasising the emotional impact.

> *“I was a little bit worried because you hear stories on the news and whatnot that some people took a turn for the worse after a week.” (Male, 60-69)*

Patient experiences due to a mandatory quarantine period (typically 2 weeks), were classified within their own theme. These impacts included being housebound, having to stay away from others, and boredom.

### Clinician interviews

Five clinicians were interviewed (n=4 in the US; n=1 in South Korea, where all COVID-19 patients are hospitalised regardless of symptom severity). The Korean clinician was informed of the focus of this study and asked to comment specifically on their experience treating mild-to-moderate cases. See Table 2 for full clinician backgrounds.

**Table 2.** Overview of clinician sample characteristics.

| **Current position(s)** | **Specialty/**<br>**training** | **No. of years**<br>**practicing** | **Experience with COVID-19 patients** |
| --- | --- | --- | --- |
| Assistant professor and attending physician at an academic medical centre | Pulmonary and critical care | 8–9 | Treating patients in person and remotely, hospitalised and non-hospitalised, pre- and post-diagnosis. |
| Attending physician in infectious diseases at an urban federally qualified health centre | Internal medicine | 10 | Treats patients in person and remotely. Sees patients with and without symptoms (patients are screened at the door to the clinic), about 10 COVID-19-positive patients per week. |
| Infectious disease department | Infectious disease | Not stated | Treating only hospitalised patients, as all COVID-19 patients are hospitalised in Korea. |
| Pulmonary critical care physician at a community teaching hospital and director of the sleep centre | Pulmonary critical care, internal medicine, and sleep medicine | 16 | Treating patients who have had COVID-19 in the past, but are not recovering (eg, retained shortness of breath or cough). Seeing 4–5 patients in the office per week. Generally performing second-line treatment only. |
| Fellow internist, private practitioner at a community hospital, and internal medicine director of a rehab hospital | Internal medicine | 25 | Treating 50/50 hospitalised and non-hospitalised patients. Sees 4–5 people per week who are concerned about COVID-19. Conducting visits remotely and in person. |

Clinicians confirmed all symptoms reported by patients and described three distinct symptomatic presentations of COVID-19: (1) fever-dominant presentation (high fevers and associated symptoms eg, headache, malaise, fatigue and muscle aches); (2) respiratory presentation (cough, shortness of breath and difficulty breathing); and (3) gastrointestinal presentation (diarrhoea, vomiting and nausea). Clinicians agreed that patients often experience symptoms across presentations during their time with COVID-19.

According to clinicians, patients often sought medical attention due to changes in taste/smell, fever, aches, weakness, cough, sore throat, joint problems, nausea, vomiting and diarrhoea. Clinicians confirmed patient reports regarding the varied nature of symptom severity and duration. Lingering symptoms were fatigue, weakness, shortness of breath, fever, joint pain, cough, loss of taste/smell and mental fatigue. Clinicians indicated that most of their patients returned to baseline health once symptom-free, but a minority took longer to return to pre-COVID-19 health due to changes in mental clarity (eg, slowed thinking, mental fatigue) or energy levels.

## Discussion

As of March 2021, there were no published qualitative studies assessing the symptoms or daily life impacts experienced by outpatients with COVID-19. The symptoms that made up the patient experience in the current study are consistent with those reported by the CDC.^9^

The symptom experience of patients with COVID-19 was collected and summarised in a clinically grounded conceptual model comprising the following groups of symptoms: upper and lower respiratory, systemic, gastrointestinal and loss of or alteration in in taste or smell. This model is a basis to determine which symptoms to consider when evaluating treatment benefit in COVID-19. We consider this model to be a comprehensive representation of COVID-19 symptoms based on patients’ heterogenous symptom profiles and the achievement of conceptual saturation, indicating the adequacy of the sample size.

Although the symptoms in the conceptual model can be considered comprehensive at a group level, the individual combination of symptoms per patient varied greatly. This was supported by clinician feedback (lack of one, clear symptomatic COVID-19 profile) and consistent with the current medical knowledge of COVID-19 manifestations. The individual patient experience also varies in terms of symptom duration, severity and timing of onset.

One key finding not described by clinicians was the heterogeneity of altered senses of smell and taste. Clinicians commonly reported that both senses would be lost completely, most often in conjunction with one another. Patients described experiencing changes in only one of these senses, or in both senses non-concurrently; patients also described alterations in these senses, rather than loss (ie, experiencing certain atypical smells and tastes). This alteration of smell or taste was commonly reported in this study, and is one of the strongest predictors of a COVID-19 infection in mild-moderate cases.^16^ Although frequent in other influenza infections, this is distinct as the way it manifests itself in COVID-19 can be present without other upper-respiratory symptoms.^17^

The daily life impacts associated with COVID-19 included ADLs; broad daily activities; leisure, social and professional activities; and emotional impacts from receiving a diagnosis/living with COVID-19. The range of impacts, both physical and emotional, played a significant role in the experience that outpatients with COVID-19 depicted. Despite the current study interviewing outpatients with mild-to-moderate cases of COVID-19 (at the time of interviewing), all patients felt that their daily lives were impacted. Given that the impacts of daily living occurred concomitantly with symptoms, we can hypothesise that a reduction in symptoms should reduce at least some impacts described by patients. Further qualitative research should aim to understand the relationship between specific symptoms of COVID-19 and associated impacts of daily living.

This study had some limitations. First, there was a time lapse between the onset of COVID-19 symptoms and the interview. Patients, on average, completed the interview 12 days after their positive diagnosis. Further studies could explore the symptom trajectory at an earlier stage of their COVID-19 experience, but challenges in recruitment made this difficult, such as the time between being tested and receiving the results, as well as restrictions on public spaces. Future studies may be able to overcome recruitment challenges with more readily available rapid testing. However, patients had vivid memories of their symptoms and provided detailed information on the experience. Second, the participants were not of a representative racial/ethnic background and therefore further research in a more diverse population is required. A final limitation was young average age; age is a documented risk factor for COVID-19 infection, and older patients have been disproportionately affected by COVID-19 symptoms.^18^ However, younger patients did report a range of symptoms and daily life impacts, emphasising the importance of infection prevention measures in younger adults.^19^ Further, the types of symptoms experienced by patients did not differ by participant age; further research should be conducted to confirm differentiation of symptoms by age groups. Additional research should also be conducted to explore the symptom experience quantitatively, including the long-term effects of COVID-19, through non-interventional observational studies and clinical trials.

This study provides unique and valuable insights from the patient perspective into the symptom experience in outpatients with COVID-19 and how these in turn could impact their lives more broadly, including physical, emotional and psychological functioning. Existing clinical guidelines for treating outpatients with mild-to-moderate COVID-19 are limited; as of 8 April 2021, the National Institute of Health provides guidelines for those at high risk of clinical progression but does not suggest treatment methods for the remainder of the outpatient population.^20^ The current study could supplement existing knowledge in informing treatment guidelines.

## Conclusion

In this qualitative research, we documented the symptomatic experience of outpatients with COVID-19 and its impacts on daily life. It provides an evidence base for selecting the symptoms and impacts important for evaluating the benefit of treatment in outpatients. While reported symptoms were in line with expectations,^5,9,21^ patients offered new insights into the experiences of smell and taste during COVID-19 infection, including descriptions of lost, declined or altered senses occurring both separately and together. Another novel finding of this study was the patient reporting of the impacts that COVID-19 symptoms had on their daily lives. Future studies should explore the symptoms and impacts of COVID-19 longitudinally, to better understand their early onset, progression/resolution, and possible long-term implications of COVID-19 (eg, lingering symptoms and daily life impacts).

## Supporting information

APodolanczuk_ICMJE Form

ARams_ICMJE Form

DRofail_ICMJE Form

KPrzydzial_ICMJE Form

NMcGale_ICMJE Form

PMarquis_ICMJE Form

SSivapalasingam_ICMJE Form

VMastey_ICMJE Form

## Data Availability

Not applicable

## Acknowledgements

We thank the study participants, their families and the clinicians involved in this trial. The authors also thank Prime, Knutsford, United Kingdom for manuscript formatting and copy-editing suggestions.

## Data sharing

Not applicable.

## Contributions

DR contributed to conceptualization, funding acquisition, methodology, project administration, supervision, visualization, and writing (original draft, review & editing).

NM contributed to conceptualization, data curation, methodology, formal analysis, project administration, supervision, visualization, and writing (review & editing).

AP contributed to conceptualization, supervision, and writing (review & editing).

AR contributed to data curation, methodology, formal analysis, visualization, and writing (original draft, review, & editing).

KP contributed to data curation, methodology, formal analysis, visualization, and writing (original draft, review, & editing).

SS contributed to conceptualization, supervision, and writing (review & editing).

VM contributed to conceptualization, supervision, and writing (review & editing).

PM contributed to conceptualization, formal analysis, supervision, visualization, and writing (review & editing).

